# Disparities in Knowledge, Attitude and Practices on Mental Health among Healthcare Workers and Community members in Meru County, Kenya

**DOI:** 10.1101/2022.03.09.22270872

**Authors:** Colleta Kiilu, Jack Musembi, Diana Mukami, Catherine Mwenda, Yvonne Opanga, George Kimathi

## Abstract

**Background:** Mental health (MH) remains a neglected priority in many low and middle-income countries. Currently, there is inadequate data on the prevalence of mental health in Kenya. This is compounded by huge inequity in the distribution of skilled human resources for mental health services. Inadequate knowledge about mental health and negative attitudes towards people with mental health disorders is widespread among the general public.

**Methods:** This was a descriptive cross-sectional survey that utilised mixed methods for data collection. A total of 535 community members and 109 healthcare workers (HCWs) were targeted for the study. All cadres of healthcare workers in the selected health facilities who voluntarily consented to participate were recruited through simple random sampling. Data were collected using household surveys, Key Informant Interviews (KIIS) with facility in-charges; Focus Group Discussions (FGD) with community members particularly community Health Volunteers (CHVs) and youth; and In-depth Interviews (IDI) with community gate keepers such as religious leaders, *Religious leader*, Chiefs/sub-Chiefs, and traditional leaders. Data analysis included simple univariate frequencies of questions chosen to reflect the key concepts on mental health. Descriptive statistics were used to determine frequencies and percentages for the different variables under study. For qualitative data, thematic analysis was applied to generate themes through deductive and inductive methods. Triangulation of qualitative and quantitative data was conducted.

**Results:** Approximately 39.1% of respondents reported to have had a family member with mental illness and 68% of HCWs reported to have diagnosed a patient with mental illness. 64% of respondents cited causes of mental disorders as witchcraft; generational curses in some families; genetic factors; drug and substance abuse especially marijuana; social and economic/financial pressures; and injuries from accidents. 93.3% of the HCWs reported to have referred patients to a mental health facility. Only 29.4% of the HCWs reported having counselling services in the facilities for patients with mental health needs. Majority (90.8% HCWs and 62.3% community members) reported that it is convenient for patients with MH needs and illness from the community to access the health care facilities and that MH services were available and mainly offered at the Meru Teaching and Referral Hospital (81.7% HCWs and 53.8% community members). Majority of HCWs (89.9%) reported that MH services were affordable to community members. On the contrary, only 44.4% of community members reported that MH services are affordable. The HCWs reported that the drugs were given for free at the health facilities while community members reported that sometimes stock-outs in drugs for MH existed in which case they would purchase drugs from pharmacies. Majority (96.4% HCWs and 62.5% community members) reported that patients with mental health^1^ needs and illness^2^ are treated with respect in the facilities. Aside from health facilities, community members also seek mental health services from: religious leaders; traditional healers including the *Religious leader* who were approached for cleansing if one believed that the mental health issues were a curse for committing certain offenses. It was also evident that some families did not seek any kind of help for their relatives with mental health illness and needs, with some even detaining them.

**Conclusion:** This study adds to the global knowledge on mental health among healthcare workers and community members providing vital data at service delivery level from an African developing country perspective. There is evidence of high burden of MH in the county with very few facilities offering MH services for patients. The existence of myths and misconceptions around the causes of MH is evident and needs to be addressed. There are also evident disparities in the perception of HCWs and Community members in MH with regards to availability and affordability of MH services and access to MH drugs. Communities still seek MH services from traditionalists and some people still neglect MH cases. Sustained poor mental health of individuals, families, the communities including healthcare workers has an enormous contribution towards negative health seeking behavior as well as social capital, an important determinant of health not just in Kenya but in many rural settings across the world. With this therefore, there is need to build the capacity of health care workers and create awareness to the community members as well as strengthen health systems to tackle MH.

## Background

Mental health is defined as “a state of well-being whereby individuals recognize and realize their abilities, are able to cope with the normal stresses of life, work productively and fruitfully, and make a contribution to their communities”(Bebbington, 2001). Positive mental health includes emotion, cognition, and social functioning and coherence(WHO, 2019). Mental health is a key determinant of overall health and socio-economic development. It influences a variety of outcomes for individuals and communities such as healthier lifestyles; better physical health; improved recovery from illness; fewer limitations in daily living; higher education attainment; greater productivity, employment and earnings; better relationships with adults and with children; more social cohesion and engagement and improved quality of life(WHO, 2019).

Mental health is a neglected priority in many low and middle-income countries(World Health Organization, 2011). There have been several recent international calls to scale up mental health service provision in these countries(Patel et al., 2008). To assist this scaling up process, there is reasonably good evidence for a range of cost-effective interventions, including interventions in African settings. Despite continuous evidence on the impact of mental disorders, there has been very limited focus in global efforts at alleviating poverty through investment in health(Kiima & Jenkins, 2010). However, a key gap is in interventions that have a comprehensive community-based approach to addressing the needs of people with mental health problems living in poverty.

The relationship between mental health and poverty in low and middle-income countries is complex and includes both social causation and social drift (or social selection) mechanisms. In relation to social causation, people living in poverty are at increased risk of developing mental health problems through the stress of living in conditions of deprivation, increased risk for trauma and other negative life events, increased obstetric risks, social exclusion and food insecurity (Tsai et al., 2012). The 65^th^ World Health Assembly (WHA) adopted Resolution WHA 65.4 on the global burden of mental disorders and the need for a comprehensive coordinated response from the health and social sectors at country level. Subsequently, during the 66^th^ World Health Assembly, Resolution WHA 66.8 was adopted which called on member states to develop comprehensive mental health action plans in line with the Global Comprehensive Mental Health Action Plan 2013-2020.

The Kenya Mental Health Policy 2015-2030 provides for a framework on interventions for securing mental health systems reforms in Kenya(MoH, 2015). This is in line with the Constitution of Kenya 2010, the Kenya Vision 2030, the Kenya Health Policy (2014-2030) and the global commitments. The Constitution of Kenya 2010 article 43 (1) (a) Provides that “every person has the right to the highest attainable standard of health, which includes the right to healthcare services” including mental health(GOK, 2010).

Data and information on the prevalence of mental health, neurological, and substance use (MNS) in Kenya is currently inadequate. It is however estimated that one in every 10 people suffer from a common mental disorder with the number increasing to one in every four people among patients attending routine outpatient services. The leading mental illnesses diagnosed in Kenya are depression and anxiety disorders followed by substance use disorders(MOH, 2020).

Meru County is located in the upper Eastern part of Kenya. The County has only one mental health unit situated in the Meru Teaching and Referral Hospital (MTRH). Thus, providing mental health services for a population of about 1,545,714 residents of Meru County as well as neighbouring counties(Government, 2018). The diagnostic and treatment gaps are made worse by the prevailing conditions of frail mental health systems as a whole. This survey’s intention was to generate the much-needed insights about the current knowledge levels, prevailing attitudes, practices and services towards mental health, care and treatment from healthcare workers and the general public. Information generated not only informed the training needs and gaps among healthcare workers, but also identified opportunities for community education, and advocacy for improved mental health support system at various levels of care.

## 2.0 Methods

A cross sectional survey design was adopted. Data was collected from community members and HCWs using a mixed method approach that is descriptive, analytical and consultative entailing triangulation of both qualitative and quantitative data to enhance validity and reliability of the research findings. Questionnaires were used to collect data on various mental health related aspects at community level while interview schedules were used to collect data on health workers’ opinions and perceptions about mental health issues. Socio-demographic characteristics were collected using a self-report instrument for variables such as age, gender, basic medical qualifications, year of training, employment status, and deployment location.

The survey was conducted in Meru County, located in the upper Eastern part of Kenya, to the East of Mt. Kenya whose peak cuts through the southern border. The survey targeted community members and health care workers in Meru County. The community members that took part in the survey included household’s heads, youth, religious leaders, herbalists/traditional healers, Religious leader, police officers, and the chiefs/sub-chiefs. Different cadres of health care workers from the selected health facilities were involved in the survey. These included Doctors, Clinical Officers, Nurses, and representatives from the Public Health department including the Community Health Extension Workers (CHEWS). The survey also included Community Health Volunteers (CHVs).

For the community at the household level, systematic sampling and simple random sampling techniques were adopted to identify respondents at every stage using a sample size proportionate to sub counties population size. In the first stage, simple random sampling technique was used to select target sub-locations from all the sub-counties as the primary sampling unit (PSU) using proportion to size method. Target survey sites were chosen using simple random methods. The selection of specific villages (as the secondary sampling unit) from the selected sub-units was done using simple random sampling. The last stage of sampling involved the application of systematic sampling method with the interval nth being determined by dividing the number of households in the respective villages. To identify the first household, purposive sampling was used to pick the first household in the direction with most households.

strict adherence to COVID-19 prevention protocols and procedures such as wearing of face masks for all survey respondents and Research Assistants, social distance and hand washing with soap and water or using sanitizers was observed. The research team was equipped with information on how and where to call in case they experience exposure or symptoms of COVID-19 or notice a potential respondent with symptoms of COVID-19 and need professional support to be referred appropriately

The Research Assistants were trained to ensure ethical conduct is clearly understood and implemented. The training included focused sessions and practical exercises on COVID-19 prevention, the informed consent process, the importance of protecting the privacy of subjects, and the confidentiality of the information obtained. Research Assistants informed respondents that they do not have to answer any question they do not wish to. The interviews were conducted during times that were convenient to the informants. Finally, all survey findings were kept anonymous and not linked to any unique identifiers with respondent confidentiality maintained throughout the survey process.

### 2.6 Data analysis

Data was collected using ODK and analysed using the Statistical Package for Social Sciences (SPSS) software version 21. This involved removing any cases that were outside the inclusion criteria, and identifying responses that were improbable (outside the normally expected range) or impossible. This entailed simple univariate frequencies of questions chosen to reflect the key concepts on mental health. Descriptive statistics were used to determine frequencies and percentages for different variables under survey. The results were presented using graphs, tables and pie charts. Qualitative data from KIIs and FGDs was transcribed then analysed by coding into similar themes using Nvivo 11 qualitative data analysis software.

## 3.0 Results

### 3.1 Socio Demographic and Other Characteristics of the Respondents

A total of 535 household respondents were included in the survey. This translates to a 100% survey response rate. The mean age of the respondents was 44.27 years. Among the total respondents, more than half (56.4%) were females and 79.1% were married. The majority (98.9%) of the respondents were Christians, while half (50.3%) of the respondents did not have formal education. Majority (90.5%) of the respondents were aware of mental health, with 35.1% reporting having a family member with mental health needs. The characteristics of the respondents are summarized in Table 1. Among the health care workers, a total of 109 health care workers were included in the survey. Their mean age was 34.36 years. More than half (57.8%) of the HCWs who participated in the survey were females, and (69.7%) were married. All (100%) the HCWs respondents were Christians and 64.2% had diploma level education. Only 32.1% of the respondents had training on mental health.

**Table 1:**
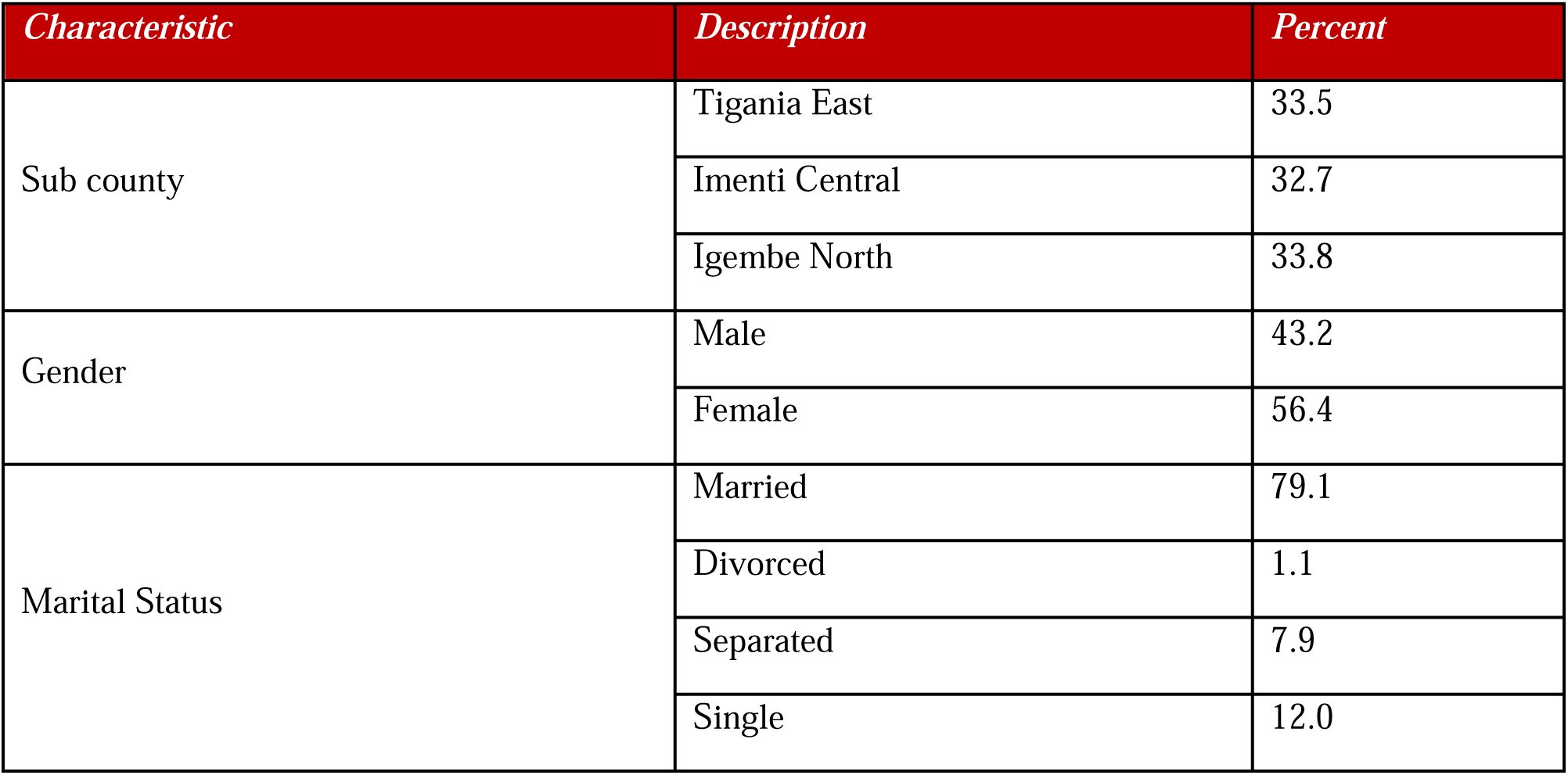
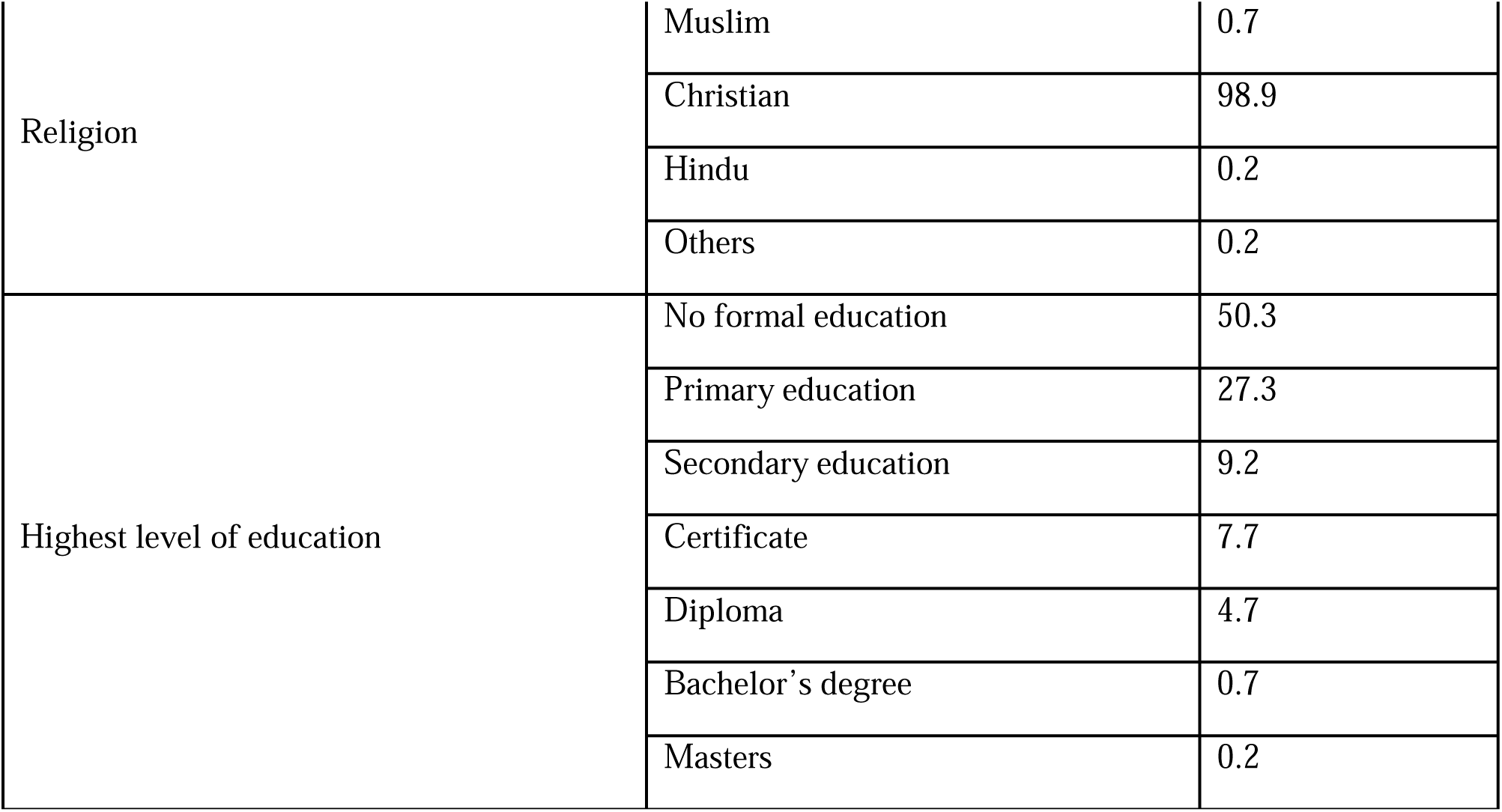
Socio-demographic Characteristics for the household respondents

| *Characteristic* | *Description* | *Percent* |
| --- | --- | --- |
| Sub county | Tigania East | 33.5 |
|  | Imenti Central | 32.7 |
|  | Igembe North | 33.8 |
| Gender | Male | 43.2 |
|  | Female | 56.4 |
| Marital Status | Married | 79.1 |
|  | Divorced | 1.1 |
|  | Separated | 7.9 |
|  | Single | 12.0 |
| Religion | Muslim | 0.7 |
|  | Christian | 98.9 |
|  | Hindu | 0.2 |
|  | Others | 0.2 |
| Highest level of education | No formal education | 50.3 |
|  | Primary education | 27.3 |
|  | Secondary education | 9.2 |
|  | Certificate | 7.7 |
|  | Diploma | 4.7 |
|  | Bachelor's degree | 0.7 |
|  | Masters | 0.2 |

### 3.2 Knowledge Levels among HCWs and Community Members on Mental Health and Illness in Meru County

#### 3.2.1 Community’s Knowledge on Mental Health Needs and Illness

The survey explored the community’s knowledge on mental health needs and illness. According to the community members, based on the qualitative data obtained from the survey, mental disorders are caused by among other things: witchcraft; generational curses in some families; genetic factors where one inherits or is born with a mental disorder; drug and substance abuse, especially marijuana; social and economic/financial pressures, and injuries from accidents which leave some people with mental disorders. Below are some excerpts on the causes of mental disorders in the community;

> *Some people are born with the condition [mental disorder] and they grow up with it. Others get mad as a result of drug and substance abuse especially marijuana*. (IDI_Respondent)
>
> *Some can be inherited; others can be due to the environment for instance if an accident happens and one is injured maybe his head to an extent that his brain is affected it can lead to mental health challenges. Still some diseases can lead to some people having those mental issues among many other reasons*. (IDI_Respondent)
>
> *Lack of financial resources contributes in mental health problems. When money lacks and one had planned to do several things for instance paying school fees, taking care of your wife etc. he or she ends up getting stressed up and eventually gets into mental health issues. So generally, poverty is a big contributor towards getting mental health issues in our communities* (R6: FGD_Participant)
>
> *People who have been bewitched, some other people have taken drugs and other substances. Others are thieves who have been bewitched for stealing. Some are born like that in communities where they have mad people while others in an effort to have quick money, they end up stealing then the owner bewitches them* (IDI_Respondent)

Witchcraft as a cause of mental disorders was also pointed in the survey where majority (64.1%) of the respondents were of the opinion that mental disorders are caused by possession of evil spirits.

Most (82.4%) of the respondents were aware that mental disorders are not contagious, with most (89.3%) reporting that mental health and illness issues are curable. Majority (91.4%) were of the opinion that mental disorders are common and can affect people of all ages and backgrounds. Despite this, it was reported that the prevalence is higher among males, and more especially the youth. Most (76.4%) of the respondents were of the opinion that people with mental disorders can have fulfilling, meaningful lives. Majority (87.5%) however reported that most people with mental disorders are dangerous.

With regards to knowledge of community members as to whether or not people from the community sought mental health services in healthcare facilities, majority (78.7%) of the respondents reported that community members sought mental health services from healthcare facilities. The survey established that aside from health facilities, community members seek mental health services from the religious leaders and traditional healers including the *Religious leader* – the council of elders – who were approached for cleansing if one believed that the mental health issues were as a result of a curse from committing certain offenses. The survey also revealed that some families did not seek any kind of help for their people with mental health and illness needs, with some even detaining them. When asked if they were aware of health facilities in the area offering mental health services, only 32% reported having knowledge of a health facility in the area managing mental needs and illness. It was reported that mental health services in the county were mainly offered at Meru Teaching and Referral Hospital.

#### 3.2.2 Health Care Workers Knowledge on Mental Health Needs and Illness

The survey explored HCWs knowledge on mental health needs and illness. According to the HCWs, based on the qualitative findings, mental health disorders are caused by among other factors genetics, disease, drug and substance abuse, and societal pressure. Reporting on the causes of mental disorders, one of the HCWs stated;

> *Basically, there are a number of causes. One is to do with genetics. We have families that are known to have issues of mental illnesses. Two, some diseases do cause mental illnesses. There are others that are as result of the abuse of certain drugs and substances. Others are as a result of trauma. There are many different causes; however, in the community people have different opinions on what could contribute to mental illnesses. People out there would think they have been bewitched. Others believe on certain superstitions* (KII_Respondent)

Most (93.6%) of the HCWs who participated in the survey were of the opinion that mental disorders are not caused by witchcraft, possession by evil spirits. They however indicated that there are people in the community who believe that mental disorders are caused by witchcraft.

Additionally, regarding the HCWs knowledge on mental health needs and illness, most (91.7%) of the respondents were aware that mental disorders are not contagious, with 54.1% reporting that most people with mental disorders are dangerous. Majority (98.2%) of the respondents were of the opinion that mental disorders are common and can affect people of all ages and backgrounds. Most (89%) of the respondents were of the opinion that people with mental disorders can have fulfilling, meaningful lives and that mental health is curable. More details are presented in table 2.

**Table 2:**
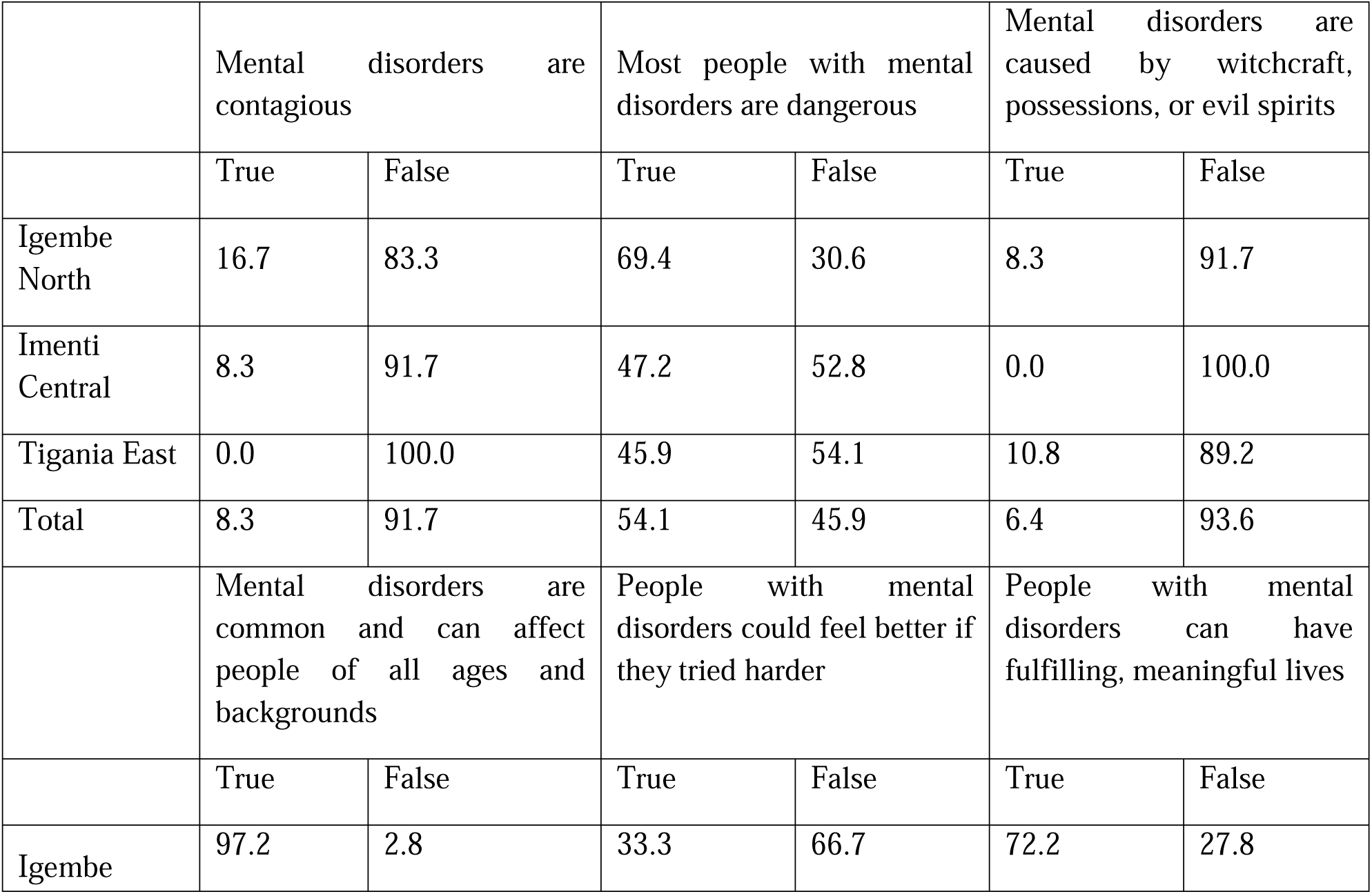
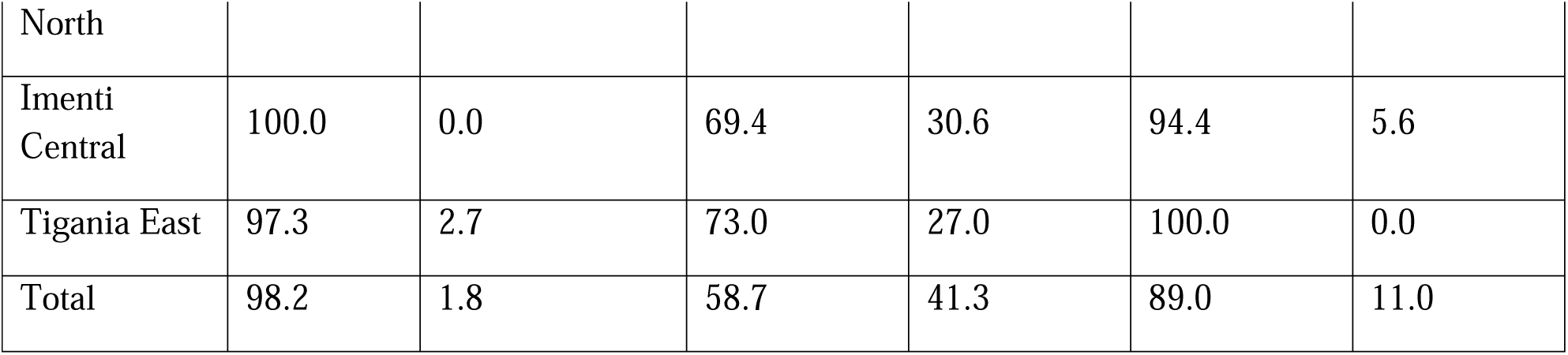
Health Care Workers Knowledge on Mental Health Needs and Illness

Ensuring consent to treatment is received from the caregiver and/or family was cited by most (60.6%) respondents as the best way of promoting respect and dignity for people with a mental condition. An assessment of the health care workers’ knowledge of depression revealed that slightly more than half (56.0%) of the respondents were aware that low energy, sleep problems, and loss of interest in usual activities best fits with an episode of depression.

Asked about a good combination treatment for depression, majority (79.8%) of the respondents cited psychosocial interventions and antidepressants as a good combination treatment for depression. This shows high levels of knowledge among health care workers on the treatment of depression.

Slightly more than half (53.2%) of the respondents were aware that people with psychosis or bipolar disorder are at high risk of stigmatization and discrimination. There were variations across the sub counties, with the least level of knowledge among health care workers being in that geographical area (33.3%).

Asked about what signs would require emergency medical treatment, slightly more than half (55%) demonstrated adequate knowledge by mentioning a seizure that lasts for more than 5 minutes, with about a quarter (22.9%) mentioning when someone starts to feel that a seizure is imminent. Details are as highlighted in figure 1.

**Figure 1:**
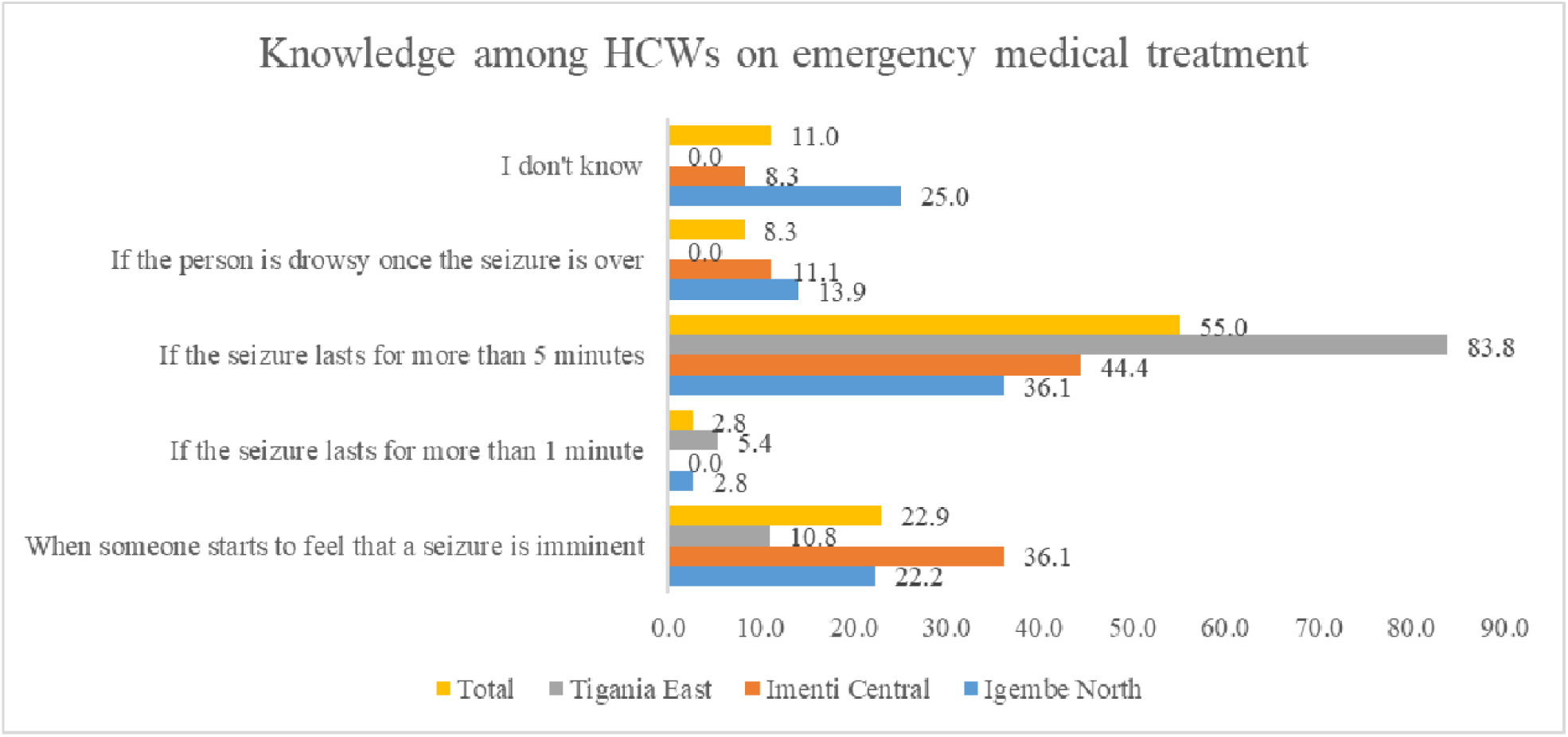
Knowledge among HCWs on emergency medical treatment

An assessment of their knowledge on child developmental disorders revealed that only 23.9% of the respondents knew that child developmental disorders is best described by attention deficit hyperactivity disorder and conduct disorder, with a nearly half (49.5%) of the respondents citing child developmental disorders involves impaired or delayed functions related to central nervous system maturation. Psychosocial intervention was reported by majority (82.6%) of the respondents as a first-line treatment for child and adolescent developmental disorders. Avoiding the use of drugs, alcohol and nicotine would be the best advice 96.3% of the respondents will give to an adolescent with a mental or behavioural disorder. Regarding presentation of dementia, a majority (69.7%) of the HCWs cited decline or problems with memory and orientation as a common presentation of dementia. There were variations across the sub counties; Imenti Central had the highest level of knowledge among HCWs (86.1%) while Tigania East had the least (51.4%).

### 3.3 Attitudes and practices of HCWs and community members towards mental health and illness in Meru County

#### 3.3.1 Attitudes of community members towards mental health and illness in Meru County

The survey explored the attitudes of community members towards mental health and illness. Based on the findings of the survey, 43.9% of the respondents were of the view that mental health needs and illnesses are as a result of a curse. There is some belief that people with mental health and illness needs may be cursed following a wrongdoing in the community, or as a result of a generational curse that is in the family. As one of the respondents reported, *“The community members blame them without even caring to know how they got in the state they are in. I have heard some members referencing the illnesses to the families of the infected. They say it’s the fault of their forefathers and other unfounded myths.”* (IDI_Respondent)

More than half (53.3%) of the respondents either disagreed or strongly disagreed with the view that mental health is untreatable with conventional medicine. This view was also shared by one of the IDIs, a Religious leader, who despite being a Christian and *Religious leader* noted that there are certain conditions that need more than prayers, and he himself would take the person with mental health and illness needs to the hospital. He stated *“I would take them to hospital. Although I am a Christian and go to church, I cannot take them to be prayed for. There are some things that need more than prayer.”* (IDI_Respondent)

With regards to the location of the facilities offering mental health services, a majority (66.3%) of the respondents either disagreed or strongly disagreed with the view that residents should not accept location of mental health facilities in their villages/neighbourhood to serve the needs of the local community, with 61.1% of the respondents further disagreeing with the view that locating mental health services in residential neighbourhoods endangers local residents. This finding was corroborated by responses from the IDIs who reported that they would be appreciative of a mental health clinic being set in the community citing its overall benefits in addressing mental and illness needs in the community. He stated; *“If such a chance would come and we get a place to take children with mental health challenges it would be such a blessing to us and the community at large.”* (IDI_Respondent)

Additionally, majority (66.3%) of the respondents either disagreed or strongly disagreed with the view that the best therapy for many people with a mental health condition is not to be part of a normal community. Also, majority (61.1%) of the respondents either disagreed or strongly disagreed with the view that residents have all to fear from people coming into their neighbourhood to obtain mental health services. Details are highlighted in table 3.

**Table 3:**
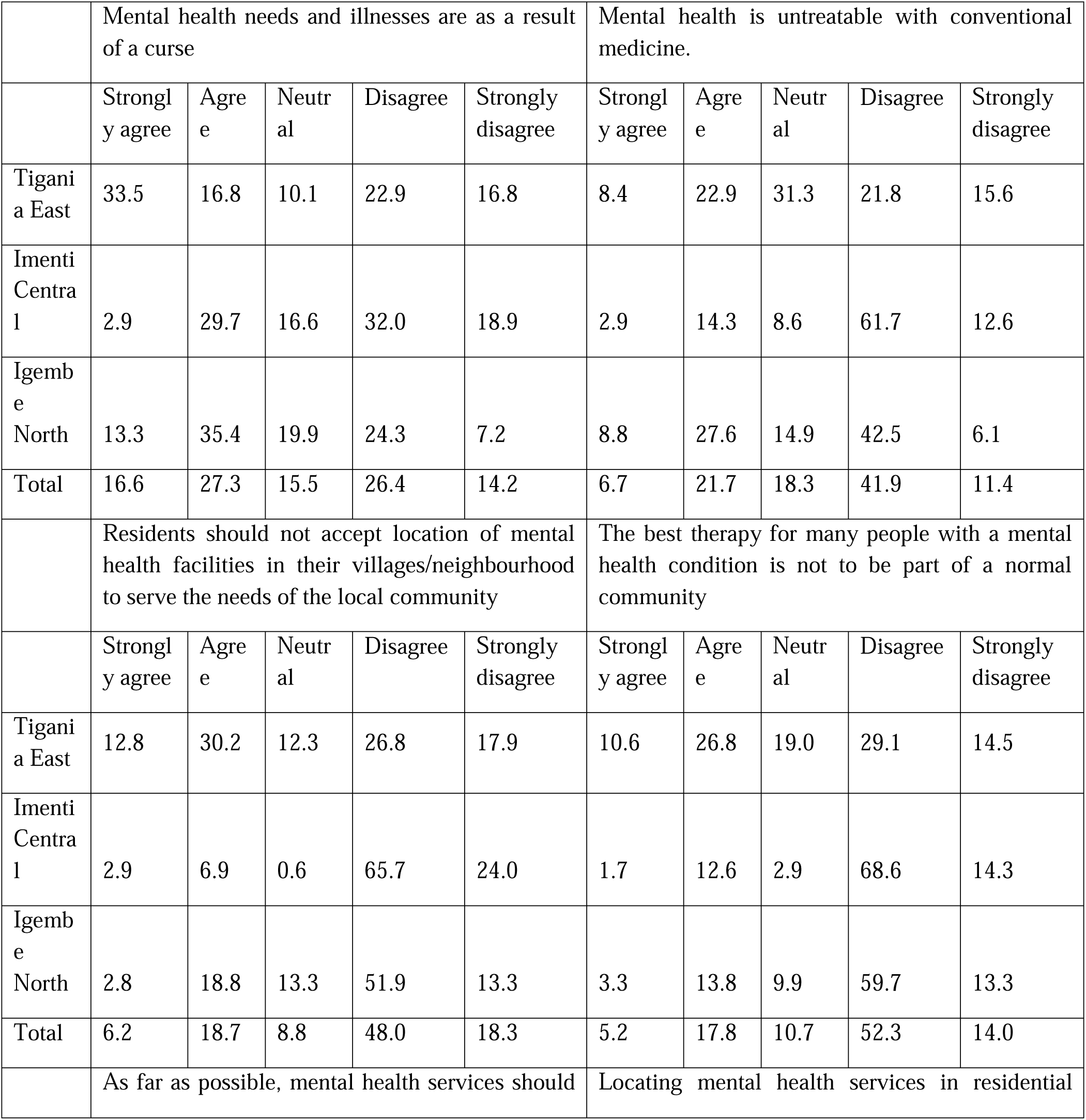
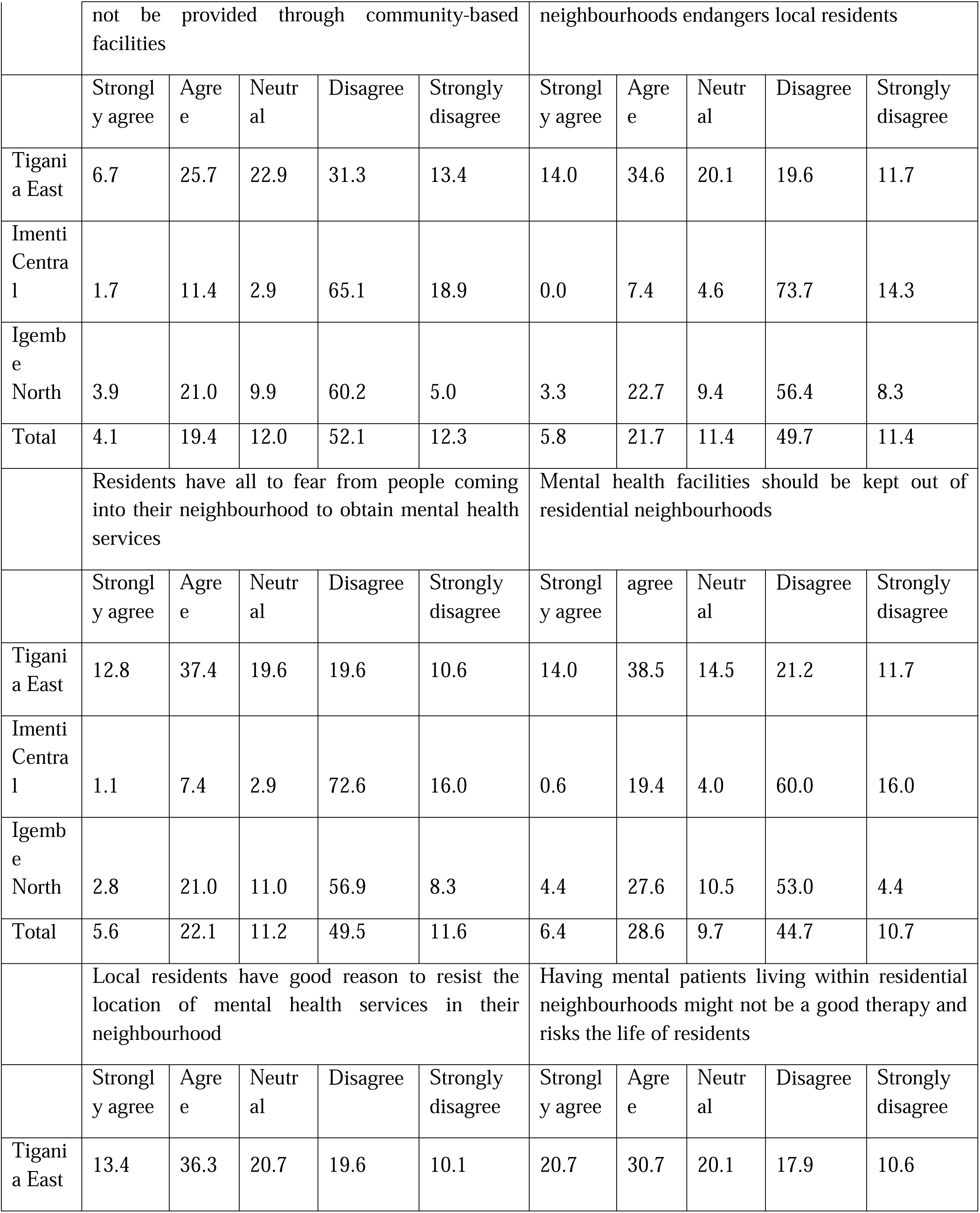
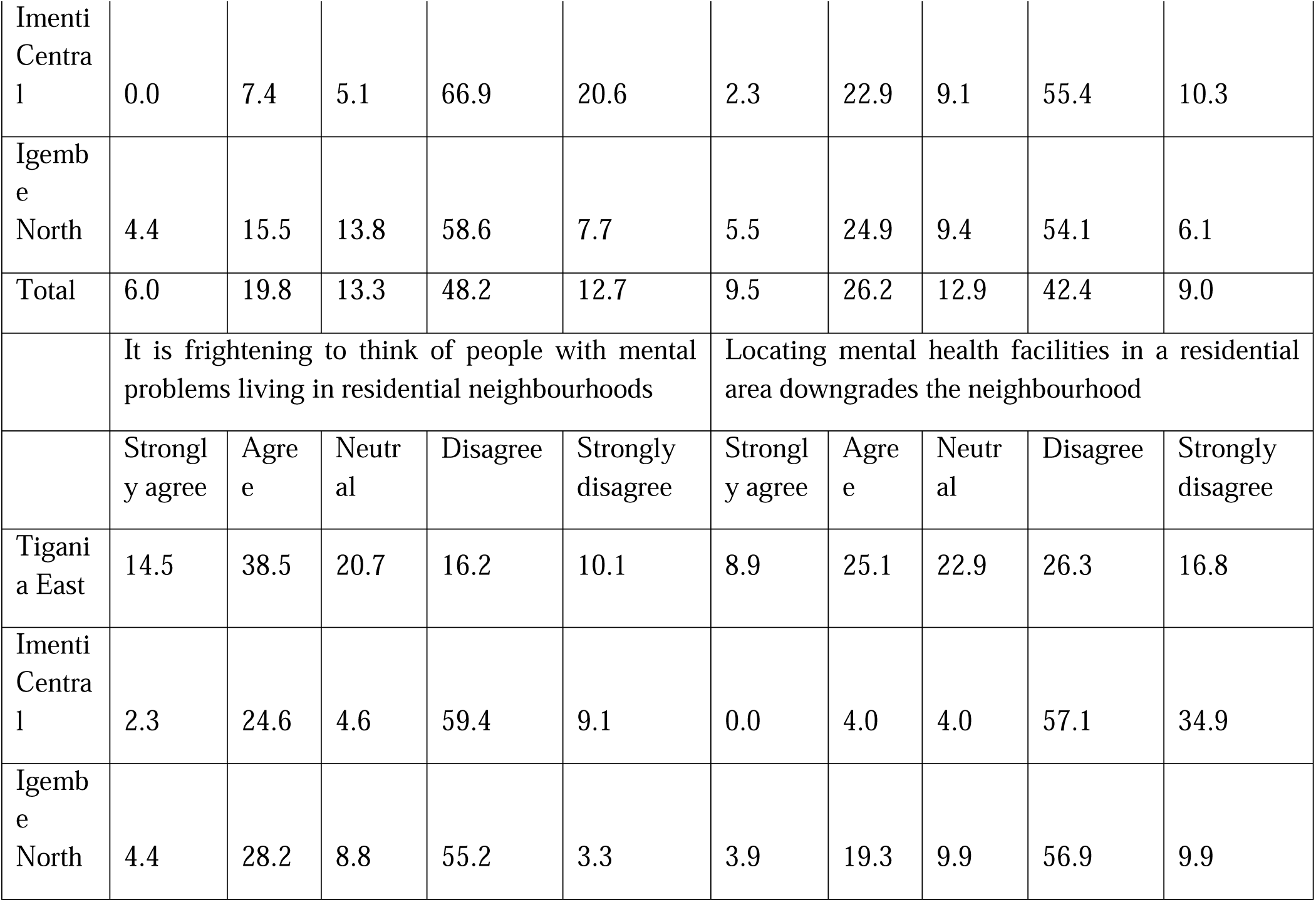
Community’s Attitude towards Mental Health Needs and Illness

|  | Mental health needs and illnesses are as a result of a curse |  |  |  |  | Mental health is untreatable with conventional medicine. |  |  |  |  |
| --- | --- | --- | --- | --- | --- | --- | --- | --- | --- | --- |
|  | Strongly agree | Agree | Neutral | Disagree | Strongly disagree | Strongly agree | Agree | Neutral | Disagree | Strongly disagree |
| Tigania East | 33.5 | 16.8 | 10.1 | 22.9 | 16.8 | 8.4 | 22.9 | 31.3 | 21.8 | 15.6 |
| Imenti Central | 2.9 | 29.7 | 16.6 | 32.0 | 18.9 | 2.9 | 14.3 | 8.6 | 61.7 | 12.6 |
| Igembe North | 13.3 | 35.4 | 19.9 | 24.3 | 7.2 | 8.8 | 27.6 | 14.9 | 42.5 | 6.1 |
| Total | 16.6 | 27.3 | 15.5 | 26.4 | 14.2 | 6.7 | 21.7 | 18.3 | 41.9 | 11.4 |
|  | Residents should not accept location of mental health facilities in their villages/neighbourhood to serve the needs of the local community |  |  |  |  | The best therapy for many people with a mental health condition is not to be part of a normal community |  |  |  |  |
|  | Strongly agree | Agree | Neutral | Disagree | Strongly disagree | Strongly agree | Agree | Neutral | Disagree | Strongly disagree |
| Tigania East | 12.8 | 30.2 | 12.3 | 26.8 | 17.9 | 10.6 | 26.8 | 19.0 | 29.1 | 14.5 |
| Imenti Central | 2.9 | 6.9 | 0.6 | 65.7 | 24.0 | 1.7 | 12.6 | 2.9 | 68.6 | 14.3 |
| Igembe North | 2.8 | 18.8 | 13.3 | 51.9 | 13.3 | 3.3 | 13.8 | 9.9 | 59.7 | 13.3 |
| Total | 6.2 | 18.7 | 8.8 | 48.0 | 18.3 | 5.2 | 17.8 | 10.7 | 52.3 | 14.0 |
|  | As far as possible, mental health services should not be provided through community-based facilities |  |  |  |  | Locating mental health services in residential neighbourhoods endangers local residents |  |  |  |  |
|  | Strongly agree | Agree | Neutral | Disagree | Strongly disagree | Strongly agree | Agree | Neutral | Disagree | Strongly disagree |
| Tigania East | 6.7 | 25.7 | 22.9 | 31.3 | 13.4 | 14.0 | 34.6 | 20.1 | 19.6 | 11.7 |
| Imenti Central | 1.7 | 11.4 | 2.9 | 65.1 | 18.9 | 0.0 | 7.4 | 4.6 | 73.7 | 14.3 |
| Igembe North | 3.9 | 21.0 | 9.9 | 60.2 | 5.0 | 3.3 | 22.7 | 9.4 | 56.4 | 8.3 |
| Total | 4.1 | 19.4 | 12.0 | 52.1 | 12.3 | 5.8 | 21.7 | 11.4 | 49.7 | 11.4 |
|  | Residents have all to fear from people coming into their neighbourhood to obtain mental health services |  |  |  |  | Mental health facilities should be kept out of residential neighbourhoods |  |  |  |  |
|  | Strongly agree | Agree | Neutral | Disagree | Strongly disagree | Strongly agree | Agree | Neutral | Disagree | Strongly disagree |
| Tigania East | 12.8 | 37.4 | 19.6 | 19.6 | 10.6 | 14.0 | 38.5 | 14.5 | 21.2 | 11.7 |
| Imenti Central | 1.1 | 7.4 | 2.9 | 72.6 | 16.0 | 0.6 | 19.4 | 4.0 | 60.0 | 16.0 |
| Igembe North | 2.8 | 21.0 | 11.0 | 56.9 | 8.3 | 4.4 | 27.6 | 10.5 | 53.0 | 4.4 |
| Total | 5.6 | 22.1 | 11.2 | 49.5 | 11.6 | 6.4 | 28.6 | 9.7 | 44.7 | 10.7 |
|  | Local residents have good reason to resist the location of mental health services in their neighbourhood |  |  |  |  | Having mental patients living within residential neighbourhoods might not be a good therapy and risks the life of residents |  |  |  |  |
|  | Strongly agree | Agree | Neutral | Disagree | Strongly disagree | Strongly agree | Agree | Neutral | Disagree | Strongly disagree |
| Tigania East | 13.4 | 36.3 | 20.7 | 19.6 | 10.1 | 20.7 | 30.7 | 20.1 | 17.9 | 10.6 |
| Imenti Central | 0.0 | 7.4 | 5.1 | 66.9 | 20.6 | 2.3 | 22.9 | 9.1 | 55.4 | 10.3 |
| Igembe North | 4.4 | 15.5 | 13.8 | 58.6 | 7.7 | 5.5 | 24.9 | 9.4 | 54.1 | 6.1 |
| Total | 6.0 | 19.8 | 13.3 | 48.2 | 12.7 | 9.5 | 26.2 | 12.9 | 42.4 | 9.0 |
|  | It is frightening to think of people with mental problems living in residential neighbourhoods |  |  |  |  | Locating mental health facilities in a residential area downgrades the neighbourhood |  |  |  |  |
|  | Strongly agree | Agree | Neutral | Disagree | Strongly disagree | Strongly agree | Agree | Neutral | Disagree | Strongly disagree |
| Tigania East | 14.5 | 38.5 | 20.7 | 16.2 | 10.1 | 8.9 | 25.1 | 22.9 | 26.3 | 16.8 |
| Imenti Central | 2.3 | 24.6 | 4.6 | 59.4 | 9.1 | 0.0 | 4.0 | 4.0 | 57.1 | 34.9 |
| Igembe North | 4.4 | 28.2 | 8.8 | 55.2 | 3.3 | 3.9 | 19.3 | 9.9 | 56.9 | 9.9 |

#### 3.3.2 Community Practices towards Mental Health Needs and Illness

To establish the community practices towards mental health and illness, the community members were asked what they would do if they had a family member with a mental health need. Majority (86.2%) reported that they would take them to a hospital. Others reported that they would take them to a spiritual/religious healer (6.7%), traditional healer (3.9%), detain them at home (1.7%), do nothing (1.3%), while others reported that they had no idea what they would do (0.2%). The results are presented in Figure 2 which were also echoed by HCWs who also reported that with cases of people with mentally ill patients, some of the community members could opt to go to the hospital; some go for prayers; some go to witchdoctors for the traditional rituals; while others would just sit back at home and do nothing. These were also reported in the IDIs. As reported by one of the respondents; *“Some go to the hospital while others come to us as herbalists. There is medication that we offer for such cases and consistent headaches. There are also those who go to be prayed for but as an individual I don’t believe in that.”* (IDI_Respondent)

**Figure 2:**
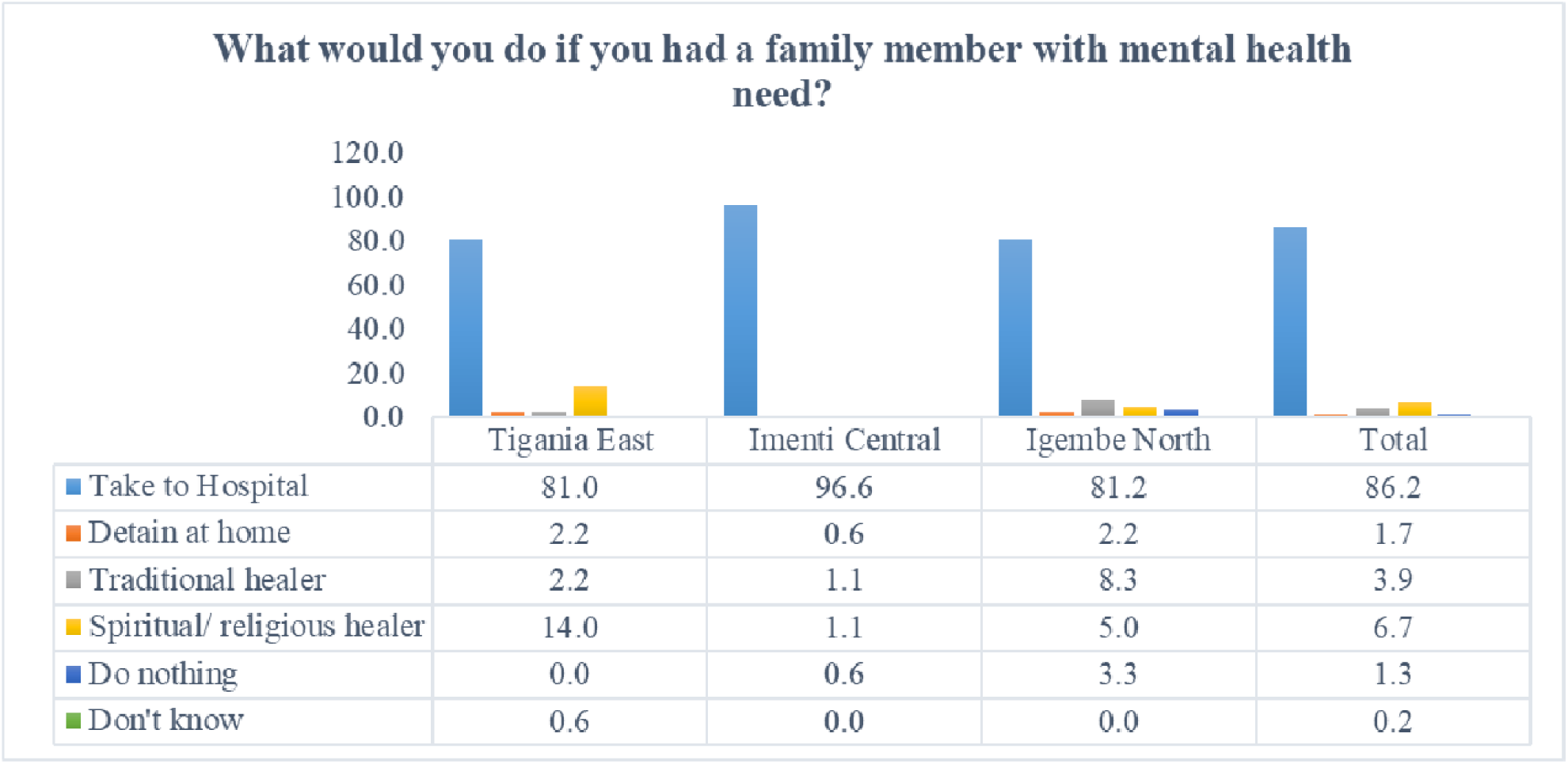
What would you do if you had a family member with mental health need?

Asked if they have ever had a family member with mental illness, 39.1% were affirmative. When asked where they sought help from, a majority (80.9%) reported to have sought help from a hospital as highlighted in figure 3. Some reported to have gone for prayers (9.6%), some did nothing (6.2%), and some detained them at home (0.5%). Others (2.9%) did not have knowledge on what was done to their family members who had mental health needs.

**Figure 3:**
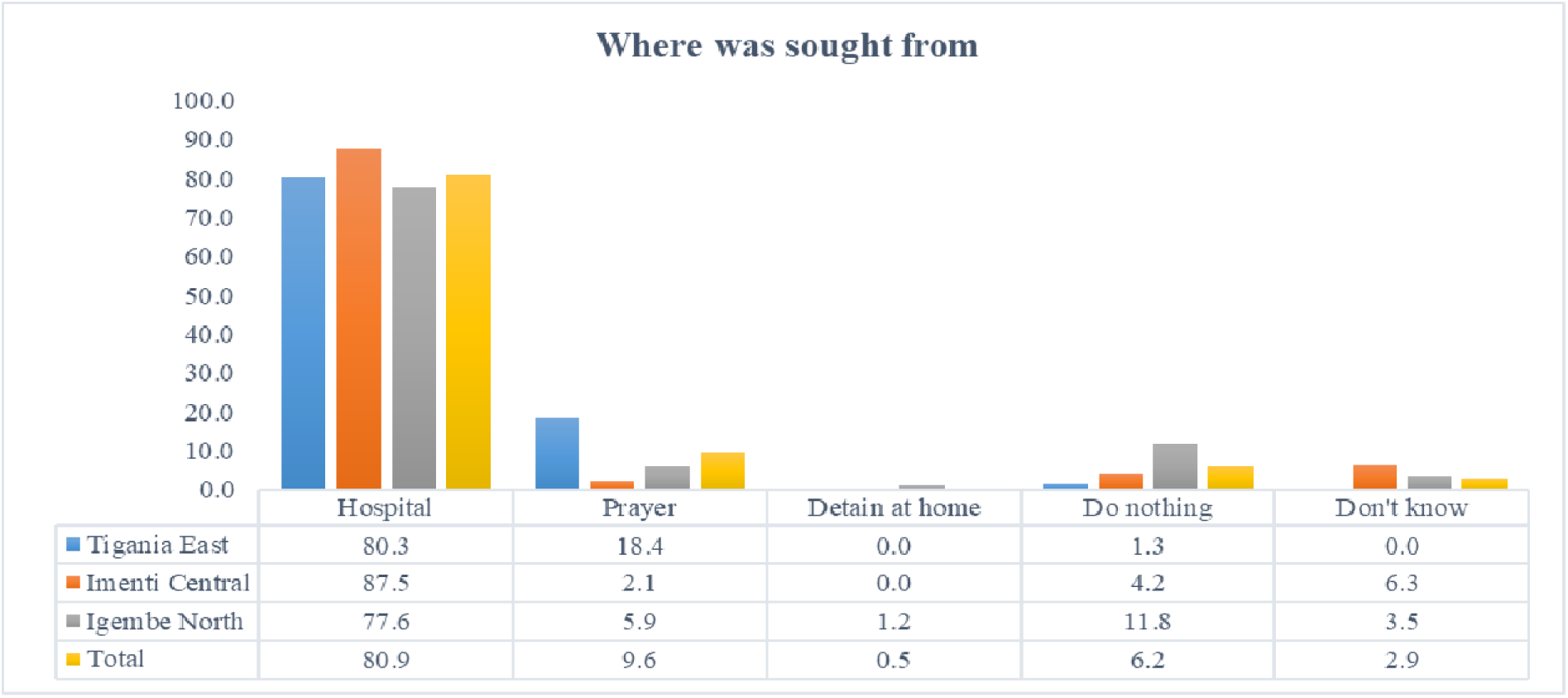
Where help was sought from

#### 3.3.3 Health Care Workers Practices towards Mental Health Needs and Illness

Regarding the practices of HCWs towards mental health needs and illness, from the interviews, it was reported that while most of HCWs may not be very competent in the treatment and management of mental illness, they were able to suspect a case of mental illness once a patient was presented after which they would refer to a Referral Hospital where there is a psychiatrist. As one of the HCWs said, *“They [the HCWS] are not very competent or very conversant with the treatment and management of mental illnesses but they are able to suspect a case of mental illness after which they refer to higher levels for management.”* (KII_Respondent)

The HCWs reported that when the patients with mental health and illness needs come, they are treated just like the other patients. However, those that come and present severe cases are treated as emergency cases and attended to immediately. Responding to this, one of the HCWs stated;

> *In the facility we treat them just like other normal patients although sometimes they come with their own issues. Some come when they are very rowdy and want to beat up everyone but generally all our patients are treated equally. We have a nurse who is at the Out-Patient Department (OPD) who does the triage, especially those cases considered emergency. The person who comes up a mental problem but is not of high degree that person is treated with other patients. Somebody rowdy and maybe tied with rope carried by two or three relatives, that one we treat as an absolute emergency and we give it first priority. When they come, they are served very fast and their referrals done very fast and referred to level 5*. (KII_Respondent)

With regards to the available services for patients with mental health and illness needs, the HCWs revealed the main place where patients with mental health and illness needs are comprehensively attended to is at the Meru Teaching and Referral Hospital which has a psychiatrist and is also well equipped. As was reported by one of the HCWs, *“The mental clinic in the General Hospital has been well equipped in terms of drugs and medical personnel. Generally basic management is done at Teaching and Referral Hospital Clinic.”* (KII_Respondent) At the lower-level facilities, the main services provided to patients with mental health and illness needs, as reported in the survey, are counselling and referrals. Other facilities, particularly the sub-county level facilities offer drug refills for those who have been diagnosed and their drugs prescribed. It was also established that some efforts were being made at increasing awareness on mental health. Reporting on the counselling and community awareness programme, one of the HCWs stated as follows;

> *I would say one is issues to do with counselling, as much as it is not necessarily targeting mental illnesses. We have conditions that are there and if not well managed and proper counselling done, they would result in severe mental illnesses. We also have extension services being done by health care workers for instance officers go to schools addressing issues to do with drug and substance abuse. These extension services work towards preventing occurrences of mental health later in life*. (KII_Respondent)

In the survey, when asked if they have ever diagnosed a patient with mental illness, 68% responded in the affirmative, with a majority (93.3%) referring the patient to a mental health facility. Only 29.4% of the respondents reported having counselling services in health facilities for patients with mental health needs.

The survey also established that only 33% conducted mental health awareness programmes in the community.

## 4.0 Discussion

The third goal of the Sustainable Development Goals as adopted by all the world governments in 2016 outlines commitments to improve mental health and improve and treat disorders associated with substance use(United Nations, n.d.). Knowledge levels on mental health are significant determinants of mental health and have the potential to improve awareness of how help and treatment for mental health patients is sought, reduce stigma against mental health illness, promote early detection of mental disorders, increase mental health outcomes and use of mental health services.(Wei et al., 2015). However, this study indicated inadequate knowledge on mental health illnesses among community members and to some extent healthcare workers. Findings for this study are in line with a study conducted in Tunisia which showed low mental health literacy levels among primary healthcare professionals. It is important to note that the level of knowledge of mental illness often affect the approach towards mentally ill people. This is often as a result of negative stereotypes and levels of stigma that lead to unwanted behaviour towards mentally ill patients(Mohamed, 2018). In Kenya, mental health illness and conditions are often misinterpreted and stigmatised. Due to lack of knowledge on mental illness and how to support individuals who are ill, most individuals always make fun of these patients, some accuse them and criticize them for the disease because of their believes on the causes of the disease. The knowledge levels as displayed by study respondents mirrors the scarcity of MH knowledge and services not just in Kenya but in the African continent. This underscores the need for increased attention to mental health by governments at all levels as well as researchers to highlight the centrality of promotive and preventative services towards MH services.

In this study, disparities in knowledge on mental illness among healthcare workers and community members was evident. This was in the context of causes of mental health, service availability, accessibility to MH services, services offered and cost of the services. For instance, only 32% reporting being aware of a health facility

On the knowledge of community members on whether or not people from the community seek mental health services from health facilities, majority (78.7%) reported that community members sought mental health services from healthcare facilities, with only 32% reporting to be aware of a health facility in the community managing mental health and illness needs. Other places where community members seek mental health services include from religious leaders and traditional healers, similar to findings from a study on access to MH services in primary healthcare facilities in Kenya(Marangu et al., 2021).

There were reported cases of other families who did not seek mental health services, with others reported to be detaining their family members with mental health and illness needs. Most Kenyans believe that mental illness is a as a result of a curse, witchcraft or evil spirits hence most relatives often lack information on how to help those affected and turn to traditional healers.(Mohamed, 2018). Mental health remains a low priority in most African countries as demonstrated by the proportion of community members who did not have adequate knowledge on mental health and illness. This was depicted by the low number of respondents with adequate knowledge on child development disorders, symptoms of substance dependence, and signs that would require emergency medical treatment. Regarding attitudes and practices of community members towards mental health and illness, majority (66.3%) of the respondents disagreed with the view that residents should not accept location of mental health facilities in their villages/neighbourhood to serve the needs of the local community, with 61.1% of the respondents further disagreeing with the view that locating mental health services in residential neighbourhoods endangers local residents. Findings from this study provide new insights into the current gaps relating to MH perceptions at community level that have a direct bearing on access and utilization of MH care and treatment services. Majority (66.3%) disagreed with the view that the best therapy for many people with a mental health condition is not to be part of a normal community. These observations are particularly relevant as mental health increases in priority not just in Kenya but also in the international agenda driving the need for prioritised mental health in Africa. These attitudes can be leveraged on in formulating community-based MH interventions to address social determinants of access and utilization of MH services in terms of community practices in relation to mental health and illness, majority (86.2%) of the respondents reported that if they had a family member with mental health need, they would take him/her to the hospital. Others reported that they would take them to a spiritual/religious healer; traditional healer; detain them at home; do nothing; while others had no idea what they would do.

Globally, 24% of countries that reported to the WHO’s 2017 mental health Atlas survey had not implemented standalone mental health policies, a proportion which rose to 46% in Africa(*M E N Ta L H E a Lt H*, n.d.). Access to health facilities that offer mental health services was pointed out as a challenge. While majority (62.3%) of the respondents reported that they had convenient access to health facilities, these facilities mainly offered counselling and referral services, with the treatment and management of mental health and illness needs primarily done at the Meru Teaching and Referral Hospital. The Meru Teaching and Referral Hospital, according to the respondents, was located far off and this meant that they incurred high costs of transport to access it. As is with most African countries, access to mental health services in Sub Saharan Africa (SSA) is largely dependent not just on availability and nature of service delivery, but also on people’s perspective regarding mental health services. This was worsened by the increased transport rates given the COVID-19 pandemic and the resulting travel restrictions. In terms of costs of services, only 44.4% of the respondents reported that the costs of mental health services were affordable. Majority reported that they sometimes had to buy some drugs in cases of stock-outs. Another area highlighted by the survey is the importance of the relationship between mental health service providers and patients where majority (54.9%) reported that the services providers had positive attitude towards patients with mental health needs with a slightly higher proportion (62.5%) of the respondents indicating that mental health patients were treated with respect. These findings concur with results from recent research using the WHO Assessment Instrument for Mental Health Systems (WHO-AIMS) pointing to lack of administrative structures for MH resulting to low prioritization of MH services access and utilization(Mutiso et al., 2020)

The survey also established that one of the challenges in access and utilization of mental health services was the lack of awareness of the availability of mental health services in facilities. The COVID-19 pandemic was also identified as an inhibiting factor in access and utilization of mental health services. The fear of contracting COVID-19 was reported to be preventing people from going to seek mental health services in health facilities.

In the case of HCWs, regarding their knowledge on mental health needs and illness, the HCWs reported that mental health and illness needs are caused by among other factors genetics, diseases, drug and substance abuse, and societal pressure. Most (93.6%) of the HCWs were of the opinion that mental disorders are not caused by witchcraft, possessions or evil spirits. Most (91.7%) of the respondents were aware that mental disorders are not contagious Majority (98.2%) of the respondents were of the opinion that mental disorders are common and can affect people of all ages and backgrounds. Most (89%) of the respondents were also of the opinion that people with mental disorders can have fulfilling, meaningful lives, with 89.9% of the respondents reporting that mental health is curable. Ensuring consent to treatment is received from the caregiver and/or family was cited by most respondents (60.6%) as the best way of promoting respect and dignity for people with a mental condition. The observed findings of positive perception are consistent with African community care ethos as described by Noor et al., that in spite of the fractured health care system in Africa, families and communities commit to support their vulnerable until appropriate care and treatment is available(Noor et al., 2006).

The survey also assessed the knowledge of the HCWs on various mental disorders including depression, acute manic episode, psychosis or bipolar disorder, child and adolescent developmental disorders, dementia among others. The survey established that while most of the HCWs were knowledgeable on a number of mental health and illness issues, majority lacked skilled training in the treatment and management of mental health and illness needs. They were able to identify a suspect mental health case and refer for attention by the psychiatrist at Meru Teaching and Referral Hospital. Asked if they have ever diagnosed a patient with mental illness, 68% responded in the affirmative with an overwhelming majority (93.3%) referring the patient to a mental health facility. While the study did not explore HCWs clinical practice treatment choices, study findings indicate that MH patients seeking care and treatment from primary healthcare facilities in rural Kenya may not receive accurate diagnosis and treatment. Only 29.4% of the respondents reported having counselling services in health facilities for patients with mental health needs. The survey established that only 33% conduct mental health awareness programmes in the community, which resonate with findings from a systematic review of evidence from low and middle-income countries focusing on Mental Health Gap Action Programme (mhGAP) (Keynejad et al., 2018)(*MhGAP*□: *Mental Health Gap Action Programme: Scaling up Care for Mental, Neurological and Substance Use Disorders*, n.d.). Survey findings point to the need for additional training towards optimal MH services provision among HCWs in Kenya. Findings from a study on exploring optimal conditions for capacity building have detailed current capabilities that can be optimized for MH capacity building(Marangu et al., 2014)

On access to and utilization of mental health services among community, majority, 90.8% of the respondents agreed that it’s convenient for patients with mental health needs and illness from the community to access the health care facility. The Meru Teaching and Referral Hospital where the clients with mental health and illness needs are referred was reported to be far thereby creating a challenge in access.

## 5.0 Conclusion

This study adds to the global knowledge on mental health among healthcare workers and community members providing vital data at service delivery level from an African developing country perspective. Based on the study findings, there is notable evidence of high burden of MH in the county with Very few facilities offer MH services like drugs and Counselling for patients. The findings also reveal the existence of myths and misconceptions around the causes of MH that need to be addressed. There are also evident disparities in the perception of HCWs and Community members in MH where although MH services are available, they seem not to be affordable for the community with Communities still seeking MH services from traditionalists and some people still neglect MH cases.

As such, it is the study’s conclusion that there potentially exists a significant opportunity for capacity building of community and healthcare workers through evidence based medical services that ultimately contribute to improved knowledge in the care, treatment and prevention of mental health disorders. The acquired knowledge consequently serves as a much-needed base for the provision of quality mental health services by both community and health workers thereby contributing to improved mental health outcomes.

In the medium term, integration of mental health training and service provision into the primary healthcare would be critical in improving the effectiveness of medical care services provided in healthcare facilities. Targeted mental health education through Continuous Medical Education (CMEs) could be mandated to motivate specific cadres of healthcare workers who are routinely charged with provision of mental health services. These messages should eventually be cascaded and availed to individuals, families, communities and society to maximize on gains form improved knowledge on subject matter

Conclusively and in view of the observed knowledge, attitude and practice gaps on the subject matter, key rapid interventions are deemed appropriate in helping bridge these gaps including: i) Mental health information to community health workers ii) A strengthened mental health support system from the County, to the Sub-County, all the way to the household level iii) Concerted awareness campaigns to address mental health issues including stigma at community level

## Data Availability

The data is available and will be shared upon request

## Appendices

## 6.0 Abbreviations

AIMs: Assessment Instrument for Mental Health Systems
CECM: County Executive Committee Member
CHEWs: Community Health Extension Workers
CHVs: Community Health Volunteers
CMEs: Continuous Medical Education
ESRC: Ethical and scientific Research Committee
FGD: Focused group Discussion
Gap: Gap Action Programme
GOK: Government of Kenya
GSK: GlaxoSmithKline
HCWs: Health Care workers
IDI: In-depth Interview
KAP: Knowledge Attitude and Practice
KII: Key Informant Interview
MH: Mental Health
MNS: Mental Health, Neurological and substance
MoH: Ministry of Health
MTRH: Meru Teaching and Referral Hospital
OCS: Officer Commanding Station
ODK: Open Data Kit
OPD: Out Patient Department
PSU: Primary Sampling Unit
SPSS: Statistical Package for Social Sciences
SSA: Sub Saharan Africa
WHA: World Health Assembly
WHO: World Health Organization

## 7.0 Declarations

### 7.1 Ethics approval and consent to participate

Ethical approval for this survey was sought from Amref Ethical and scientific Research Committee (ESRC) (ESRC P907/2020) prior to data collection. Thereafter, the research team sought approval of the survey from the MoH at both national and county levels to access health facilities. Researchers also sought permission from the facility In-charges after approval from the county in order to begin data collection exercise. The investigators sought consent from the respondents prior to commencing the data collection process. Each respondent was carefully taken through the informed consent process which included full disclosure about the survey. This also included information on the purpose of the survey, the survey procedures, the rights not to answer any question (s) or stop the interview without penalty, confidentiality, voluntariness in participation in survey, risks and benefits of the survey. No invasive procedures were used and no incentives were offered for participation. The respondents received comprehensive explanation that the survey results would be of benefit to the general practice of healthcare. Confidentiality of the data was maintained throughout the survey period and all ethical standards maintained throughout the survey period.

### 7.2 Consent for publication

The authors unanimously consent for the information in this manuscript to be considered by your journal for publication and confirm that the results in this manuscript have not been published elsewhere, nor are they under consideration by another publisher.

### 7.3 Availability of data and materials

All data generated and analysed during this survey is included in this manuscript and has been attached as a supplementary file. The dataset is also available from the corresponding author on request.

### 7.6 Authors’ contributions

CK, JM, CM and DM conceptualized, designed, performed statistical analysis and participated in the initial draft of the study. CK, JM, CK and DM participated in the design of the study and contributed to the finalization of the manuscript. The manuscript was reviewed by YO and GK with commentaries. All authors read and approved the final manuscript.

## Acknowledgments

This survey and manuscript would not have been possible without the financial support provided by GSK (GlaxoSmithKline) and the technical guidance of promotive health consultants and Meru county

## 7.8 Authors’ information

a. Colleta Kiilu; (corresponding author), Amref Health Africa
b. Diana Mukami; Amref Health Africa
c. Yvonne Opanga; Amref Health Africa
d. Catherine Mwenda; Amref Health Africa
e. Jackson Musembi; Amref Health Africa
f. George Kimathi; Amref Health Africa

## Footnotes

1 Mental health is defined as “a state of well-being whereby individuals recognize and realize their abilities, are able to cope with the normal stresses of life, work productively and fruitfully, and make a contribution to their communities” (WHO: 2003)

2 Mental illnesses are health conditions involving changes in emotion, thinking or behavior (or a combination of these). Mental illnesses are associated with distress and/or dysfunction in social, occupational or family set ups

